# Financial toxicity among cancer patients, survivors, and their families in the United Kingdom: a scoping review

**DOI:** 10.1101/2022.10.11.22280921

**Authors:** Tran Thu Ngan, Tran Hoang Tien, Michael Donnelly, Ciaran O’Neill

## Abstract

**Background:** The aim of this scoping review was to identify key research gaps and priorities in order to advance policy and practice for people living with cancer in the UK.

**Methods:** The review adhered to PRISMA guidelines for scoping review. We searched MEDLINE, EMBASE, Scopus, Web of Science, and Google Scholar on July 16, 2022. There were no restrictions in terms of study design and publication time; grey literature was included. The key words, ‘financial’ or ‘economic’, were combined with each of the following words ‘hardship/stress/burden/distress/strain/toxicity/catastrophe/consequence/impact.’

**Results:** 29/629 studies/reports published during 1982-2022 were eligible to be included in the review. No study conducted a comprehensive inquiry and reported all aspects of FT or used a validated measure of FT. The most three commonly reported outcomes related to financial hardship were financial well-being (24/29), benefit/welfare (17/29), and mental health status (16/29).

**Conclusions:** It is evident that FT is experienced by UK cancer patients/survivors and that the issue is under-researched. There is an urgent need for further research including rigorous studies which contribute to a comprehensive understanding about the nature and extent of FT, disparities in experience, the impacts of FT on outcomes, and potential solutions to alleviate FT and related problems.

## BACKGROUND

The term ‘financial toxicity’, while lacking an agreed definition, is commonly used to refer to both the objective financial burden and the subjective financial distress experienced by cancer patients, survivors, and their families as a result of cancer diagnosis and treatment [1, 2]. Objective financial burden stems from out-of-pocket payment (OOP) related to direct medical costs, direct non-medical costs (e.g., fuels for transportation, heating, special foods), and indirect non-medical costs (e.g., loss of income) [3]. Subjective financial distress, which is much more complex to assess, results from (i) the accumulation of OOP spending (direct from income or indirect through using savings or selling property); (ii) the concerns about the costs and how to deal with them; and (iii) the challenge of changing behaviours and carrying out cost coping strategies (e.g., seeking financial assistance, reduce leisure activities) [3, 4].

Financial toxicity (FT) leads to a range of adverse financial, medical, and social outcomes. Firstly and most obviously, the financial well-being of patients, survivors, and their families may be negatively impacted by FT as they may lose savings and/or assets; have lower income and slower career development due to employment disruption during the cancer treatment; accumulate debts on credit cards; and fall behind on mortgage payments [3, 5]. Regarding health outcomes, lower health-related quality of life (HRQoL) has been mentioned by several studies on the topic [4–7]. Financial toxicity may also result in additional mental health distress and conditions such as depression and anxiety - the risk of developing these kinds of mental health problems is 3 times higher among cancer survivors who experience FT compared to cancer survivors without financial hardship [8].

The risk of FT and the level of severity varies between countries. In part this can be explained by variations in access to publicly funded care and welfare supports. It might be reasonable to expect that FT is more relevant in low- and middle-income countries (LMICs) where patients are more financially vulnerable and OOP accounts for 30-50% of health care financing [9]. In fact, even in high income countries (HICs) with publicly funded health system and universal coverage, FT exists and has now become a serious issue [8, 10, 11]. Studies from Canada, Italy, Germany, and Japan have reported significant prevalence of FT among cancer patients/survivors as well as its impact on health outcomes [7, 11–13]. For example, in Italy where services are provided free at point of use by the state, a study with pooled data from 16 prospective multicentre trials reported 22.5% patients experienced FT that was significantly associated with an increased risk of death (HR 1.20, 95% CI 1.05-1.37, P = 0.007) [7].

Media reports and voluntary sector bodies report the existence of FT among cancer patients in the UK and growing concerns regarding its effects in light of rapid increases in energy prices, rising inflation and interest rates. While the overwhelming and increasing cost of treatment, patient visits, and prescriptions are covered by the government, all other direct non-medical and indirect costs still fall on the patients. Research has shown that individuals from the most socioeconomically disadvantaged groups such as lower income families, rural dwellers, minority groups, immigrants, and young people are at greater risk of financial hardship [5, 8]. Despite the importance of the problem and growing interest, there is uncertainty about the nature and extent of FT studies in the UK.

This scoping review was conducted to review available published and grey literature about FT among cancer patients, survivors, and their families in UK. The aims were to chart available empirical data about the topic of FT, identify the research gaps and key research priorities to advance policy & practice for people living with cancer in the UK.

## METHODS

The conduct of this scoping review followed the methodological framework proposed by Arksey & O’Malley (2005) [14] and Levac et al (2010) [15] as well as the PRISMA guidance for the conduct and reporting of scoping reviews [16] (See Appendix 1, Supplementary information for PRISMA-ScR checklist). There are five key stages to conducting a scoping review (plus optional stage 6).

### Stage 1: Identifying the research question

Our research question was, ‘What is known from existing literature about financial toxicity among cancer patients, survivors, and their families in the UK?’. We used the framework of FT proposed by Witte et al (2019) [4] wherein subjective financial distress was classified into three domains: (i) material conditions (e.g., the use of active and passive financial resources), (ii) psychological response (e.g., worries and concerns about their financial situation), and (iii) coping behaviours (to manage increased expenses). We decided to search for studies that reported data related to any aspect of these domains in order to ensure the breadth of coverage.

### Stage 2: Identifying relevant studies

We performed the search in four bibliographic databases including MEDLINE (via Ovid, 1946-present), EMBASE (via Ovid, 1947-present), Scopus (2004-present), and Web of Science (1900-present) on July 16, 2022. Search inquiries did not apply a time limit or restrict any study type, but an English language-only restriction was applied. The results from initial searching indicated that the term ‘financial toxicity’ was not used commonly in the UK; therefore, we applied a wide range of alternative terms and a broad encompassing search strategy. The terms, ‘financial’ and ‘economic’ respectively were combined with hardship or stress or burden or distress or strain or toxicity or catastrophic or consequence or impact. Detailed database search strategies are presented in Table S1, Supplementary information. In order to capture a wider range of study designs as well as grey literature, we searched Google Scholar and websites of relevant charity organisations including Macmillan Cancer Support, Cancer Now, Cancer Action, and Young Lives vs Cancer (formerly CLIC Sargent). Additional potential papers were retrieved from the reference lists of included studies. Literature for which full text was not available (e.g., conference abstract) were excluded as information provided in an abstract is not enough to capture the full scope of an article and hinder the accuracy and quality of interpretation.

### Stage 3: Study selection

Selection criteria for studies were based on the PEO framework (PEO – **P**opulation, **E**xposure, and **O**utcome) as follows: (1) **P**opulation: cancer patients (those who are under treatment, including patients with initial treatment after diagnosis and people with on-going treatment for metastatic cancer), cancer survivors (those who finished initial treatment), and family members of cancer patients/survivors (whether or not they were providing informal care); (2) **E**xposure: financial toxicity (including objective financial burden and subjective financial distress) experienced by the population of interest; (3) **O**utcome including financial well-being, HRQoL, mental health status and conditions such as depression and anxiety, benefits/welfare, counselling service, and any other support with a purpose that was to ease FT. Details of inclusion/exclusion criteria are presented in Table S2, Supplementary information.

All citations resulting from the searches were imported into Covidence (web-based software platform that streamlines parts of the systematic review process). After removing duplicated citations, a selection process was conducted in two steps including 1) Title and abstract screening, and 2) Full-text review. Two reviewers (TTN and THT) independently conducted these two steps. In step 1, studies were moved to full-text review if at least one reviewer voted ‘included’. In step 2, when disagreement on study inclusion occurred, final inclusion was reached by consensus.

### Stage 4: Charting the data

A data charting form was developed and piloted by the research team using three randomly selected included studies and refined accordingly (See Appendix 3, Supplementary information). Two reviewers (TTN and THT) independently extracted data. Recorded information revolved around the PEO framework and the Witte et al (2019) conceptualisation of FT [4] and included (i) General information (author(s) and their affiliation, year and type of publication, geographic coverage); (ii) Methods and participants (Objectives/research questions, study design, studied population); (iii) Exposure (FT) and outcomes (terms were used to describe the problem of FT, exposure definition or description, tools were used to measure it, outcomes of FT were studied); and (iv) Key findings.

### Stage 5: Collating, summarising, and reporting the results

We provide a descriptive numerical summary analysis of the extent, nature, and distribution of the included studies to show the dominant areas of research. We then provide a qualitative thematic analysis in which findings from included reviews were organised and presented by different outcomes of FT. Finally, the implication of findings, the broader context and recommendations regarding future research are presented.

## RESULTS

We identified 740 citations from systematic searches on four databases and included the first 200 search results on Google Scholar. After removing duplicates, 623 citations were screened by title and abstract. 53 citations were moved to full-text review, of which, 23 were included [17–39] (See Appendix 2, Supplementary information for full list of excluded reviews and justification for the exclusions). There were 6 additional studies (2 peer-reviewed articles [40, 41] and 4 grey literature reports [42–45] identified through manual searches of the reference lists of included citations and websites of relevant organizations. As a result, a total of 29 studies were included in analysis [17–45] (Figure 1).

**Figure 1:**
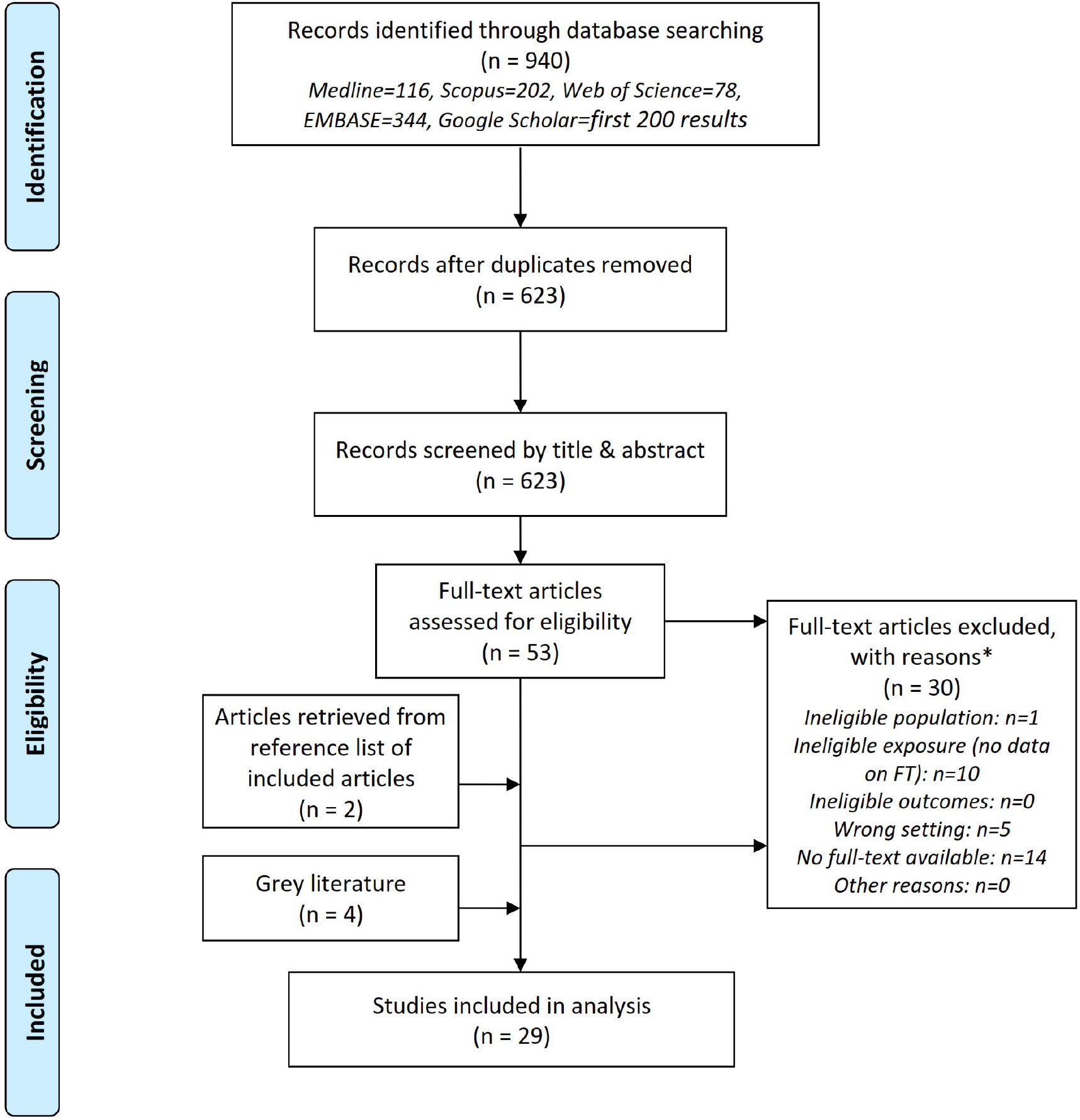
PRISMA flow diagram of literature search and selection. **\* Notes on inclusion criteria** • Population: Cancer patients (those who are under treatment), cancer survivors (those who finished initial/hospital treatment, and family members of cancer patients/survivors • Exposure (the issue): Financial toxicity (refers to both the objective financial burden and subjective financial distress) • Outcomes: Primary main outcomes are financial we/I-being, health-related quality of life, addition mental health conditions such as depression and anxiety. Secondary main outcomes are benefit/we/fare, counselling, and any other support with purpose to ease financial toxicity • Setting: United Kingdom (the whole UK or any of these following: England, Scotland, Wales, and Northern Ireland)

### Extent, nature, and distribution of studies

There was only one study [31] that had as an explicit research objective, to investigate financial toxicity. The focus of all other studies varied from the cost of cancer and its impact on family income and/or financial wellbeing to factors that influenced decisions about returning to work after treatment; from information/supportive needs of cancer patients to benefits/allowances that families were entitled to claim; from the general health and wellbeing of cancer survivors to the concerns/worries of cancer patients. No study reported all aspects of FT (objective financial burden and subjective financial distress).

Table 1 presents the numerical summary of the general information, methods, and participants of 29 included studies. The majority of the studies (21/29 or 72%) were published between 2011-2021 [20–25, 27–32, 35, 36, 38–40, 42–45] and from authors in academia (22/29 or 76%) [18–27, 29–34, 36–41]. Regarding geographic coverage, all but one study [25] (97%) included English data and 10 studies (35%) reported UK-wide data [18, 20, 28, 29, 36, 41–45]. 24 were peer-reviewed articles [17–27, 29–41] and 5 were grey literature (in the form of reports from charity organisations [42–45] and commentary from a freelance writer and welfare rights adviser [28]).

**Table 1.** Overview of general information, methods, and participants of 29 included studies.

| **Category** | **Sub-category** | **n** | **%** |
| --- | --- | --- | --- |
| **Year of publication** | Before 2001 | 1 | 3 |
|  | 2001-2010 | 7 | 24 |
|  | 2011-2022 | 21 | 72 |
| **Type of publication** | Peer-reviewed article | 24 | 83 |
|  | Grey literature | 5 | 17 |
| **Geographic coverage** | UK-Wide | 10 | 34 |
|  | Great Britain | 1 | 3 |
|  | England | 14 | 48 |
|  | England & Scotland | 1 | 3 |
|  | England & Wales | 2 | 7 |
|  | Northern Ireland | 1 | 3 |
| **Author(s)'s affiliations\*** | Academia | 22 | 76 |
|  | Charity | 5 | 17 |
|  | Hospital | 6 | 21 |
|  | Others | 1 | 3 |
| **Study design** | Mixed methods | 5 | 17 |
|  | Quantitative data (cross-sectional survey or secondary data analysis) | 8 | 28 |
|  | Qualitative data | 7 | 24 |
|  | Systematic review/review | 7 | 24 |
|  | Others | 2 | 7 |
| **Studied population\*** | Patients | 15 | 52 |
|  | Survivors | 7 | 24 |
|  | Carer/family members | 7 | 24 |
| **Sample size (quantitative)** | <200 | 5 | 38 |
|  | 201-500 | 5 | 38 |
|  | 500+ | 3 | 23 |
| **Sample size (qualitative)** | <25 | 6 | 50 |
|  | 25-60 | 5 | 42 |
|  | 60+ | 1 | 8 |

\* Not mutually exclusive

### Design, participants, exposure, and outcomes

7 (24%) papers were systematic/narrative reviews with global coverage that included the UK [20, 27, 30, 32, 33, 36, 39], 7 (24%) were qualitative research [18, 19, 24, 26, 34, 37, 40], 8 (28%) provided quantitative data from cross-sectional surveys or secondary data analysis [17, 22, 23, 25, 29, 31, 38, 41], and 5 (17%) were mixed methods studies [21, 35, 43–45]. Patients were the most studied population (in 15/29 or 52% studies) [18, 19, 21–24, 31, 35, 37, 38, 40, 42–45], followed by survivors and carer/family members (in 7/29 or 24% studies). A majority of qualitative studies had less than 60 interviews (median was 26). A majority of studies with quantitative data had a sample size smaller than 500 (median was 350). Three studies with biggest sample size of 780 [29], 1174 [21], and 1600 [43] had involved voluntary organisations (i.e., Macmillan Cancer Support) in recruitment process.

Authors used a wide range of wordings to describe the problem of financial toxicity (Figure 2). The three most common terms were ‘burden’ (n=23), hardship (n=21), and impact (n=20); followed by ‘difficulties’ (n=15), ‘problems’ (n=15), and ‘concerns’ (n=14).

**Figure 2:**
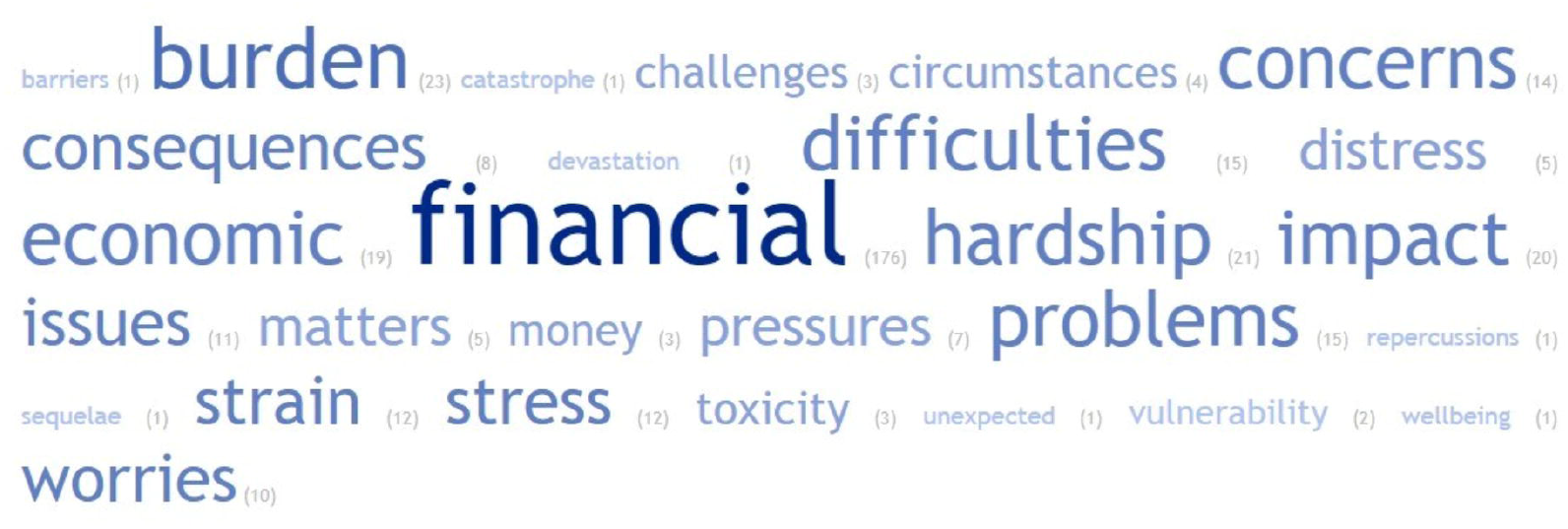
Terms were used to describe the problem of financial toxicity. **Note:** In each included studies, all term(s) used to describe ‘financial toxicity’ were listed. 29 lists of terms were then input into a tool to generate the word cloud. The number in the bracket next to each word represent the frequency that word appeared in the lists of terms. The word cloud does not show specific combination of words. In principle, any combination between either economic or financial with a noun is possible.

In the only study that specifically investigated financial toxicity, patients were surveyed and classified as facing FT when they experienced greater financial burden at follow-up compared to their assessment at baseline [31]. Financial burden was identified based on only one question (Q28) in the European Organisation for Research and Treatment of Cancer (EORTC) core Quality of Life questionnaire (EORTC QLQ-C30) which asked patients to score financial difficulty relating to disease or treatment from 1 (not at all) to 4 (very much) [31].

No study used a validated instrument to assess FT such as ‘COST - The COmprehensive Score for financial Toxicity’ [46]. Most studies used bespoke questionnaire while some used generic instruments which cover a wide range of aspects related to cancer care such as Supportive Care Needs Survey-Short Form 34 (SCNS-SF34) [38], EORTC QLQ-C30 [22, 27, 31], and Social Difficulties Inventory (SDI) [22, 27].

The most common outcomes related to financial hardship reported in included studies were financial well-being/situation (24/29 or 83%) [17, 19, 20, 22–24, 26–33, 35, 36, 38–45], benefit/welfare (17/29 or 59%) [17–19, 21–24, 26–28, 30, 32, 33, 35, 36, 41, 45], mental health (16/29 or 55%) [19–21, 24, 25, 29, 30, 35–37, 39–42, 44, 45], employment after treatment (5/29 or 17%) [26, 30, 34, 39, 40], and HRQoL (2/29 or 7%) [23, 42].

### Key findings of studies

Table 2 and 3 provide summaries of included studies’ key findings. These findings are organised thematically into six following outcome-related themes

**Table 2:** Summaries of included studies’ key findings (studies published before 2011)

| Author<br>Year | Objectives (aspects investigated) | Study<br>design | Pop | Outcomes |  |  |  |  | Key findings: (a) Financial well-being, (b) Benefits/Welfare, (c) Mental health, (d) Employment, (e) Health-related quality of life (HRQoL) |
| --- | --- | --- | --- | --- | --- | --- | --- | --- | --- |
|  |  |  |  | a | b | c | d | e |  |
| Bodkin<br>1982 [17] | Financial problems and hardship | Quan | C | x | x |  |  |  | a. Severe financial problems due to increased expenditure and loss of income<br>b. Many families received financial help towards travel, special food, and heating from charitable sources. Most did not qualify for State benefits |
| Rozmovits<br>2003 [37] | Information needs | Qual | P |  |  | x |  |  | c. Worries about loss of income |
| Chapple<br>2004 [18] | Financial concerns, perceptions, and experiences with lung cancer | Qual | P |  | x |  |  |  | b. Unaware of financial benefits or lack of information on how to claim one. Stigma in claiming financial help. |
| Eiser<br>2006 [40] | Costs of caring for a child with cancer; impact on parents' income and the contribution of government benefits and charities | Quan | C | x | x | x |  |  | a. Changes in employment impacted negatively on finances of 42.7% families. Parents were forced to give up paid employment (34.7% mothers & 1.7% fathers), reduce working hours (28.7% mothers and 37.3% fathers), or changed employment (2% mothers and 1.7% fathers)<br>b. Benefits were not received timely<br>c. 68.3% families were worried about money. Lone parents had more financial concerns than parents who were married/cohabiting |
| Hanratty<br>2007 [33] | Existence and consequences of financial stress and strain at the end of life for people dying with cancer | Sys<br>rev |  | x | x |  |  |  | a. 16% to 80% claimed that they need more financial help. Differences among sociodemographic groups: 32% working class vs 16% middle class; 80% of the black carers vs 26% of the white carers<br>b. 26% to 55% received attendance allowance |
| Kennedy<br>2007 [34] | Factors that influence decisions to return to work and the experience of returning to work for cancer survivors | Qual | S |  |  |  | x |  | d. Primary reason (50%) for returning to work is financial pressure of being off work |
| Amir<br>2008 [26] | How people have returned to the world of work | Qual | S | x | x |  | x |  | a. Built up significant debts on credit cards or fell behind with mortgage payments<br>b. Dissatisfy with the financial protection for sick workers in the social welfare context of the UK<br>c. Return to work earlier due to acute financial pressure |
|  |  |  |  | a | b | c | d | e |  |
| Moffatt<br>2010 [19] | Impact of a welfare rights advice service specifically designed for people affected by cancer and their carers in County Durham, Northeast England (UK) | Qual | P<br>+<br>C | x | x | x |  |  | a. Most of the participants experienced financial strain following their cancer diagnosis. No financial impact was reported from households where a working partner earned a high income and/or the individual was well covered by private health insurance and/or mortgage protection. Financial impact was more severe for those of working age, especially those were self-employed<br>b. Successful benefit claims was used to offset additional costs associated with cancer and lessen the impact of loss of earnings. Main barrier to access benefits was lack of knowledge about benefit entitlements<br>c. Additional stress due to money worry. Quoted: "if you've got money worries it brings you down a little bit further", "It's a hard enough worry cancer itself, without having to worry about money as well" |

*Study design – Mixed: Mixed methods, Qual: Qualitative research, Quan: Quantitative data, Rev: Narrative review, Sys rev: Systematic review*
*Pop: Studies population – C: Carers/family members, P: Patients, S: Survivors*

**Table 3:** Summaries of included studies’ key findings (studies published after 2010)

| Author<br>Year | Objectives (aspects investigated) | Study<br>design | Pop | Outcomes |  |  |  |  | Key findings: (a) Financial well-being, (b) Benefits/Welfare, (c) Mental health, (d) Employment, (e) Health-related quality of life (HRQoL) |
| --- | --- | --- | --- | --- | --- | --- | --- | --- | --- |
|  |  |  |  | a | b | c | d | e |  |
| Brooks<br>2011 [20] | Additional expenses related to cancer | Rev |  | x |  | x |  |  | a+c. Money worries increased for 68.3% families after diagnosis. Lone parents more likely to report money worries |
| Elliott<br>2011 [29] | Self-reported health and well-being | Quan | S | x |  | x |  |  | a. 15-18% of cancer survivors were in debt but not worried about it<br>a+c. 12-14% of cancer survivors with were in debt and worried about it |
| Young<br>Lives vs<br>Cancer<br>2011 [44] | Additional costs facing families, how a cancer diagnosis disrupts their employment and ability to earn income, what financial support is available, and how families cope with these various impacts | Mixed | P<br>+<br>S<br>+<br>C | x |  | x |  |  | a+c. The number of parents who said that money was 'often' or 'frequently' a worry increased eight-fold after diagnosis, from 8% to 65%. 76% families said that childhood cancer had been a 'big problem' for their finances. |
| Amir<br>2012 [40] | Effects of cancer's related financial hardship/worries on family life (i.e., financial concerns of people affected by cancer) | Qual | P<br>+<br>C | x |  | x | x |  | a. Loss of income, especially for patients were in paid employment or self-employed at the time of diagnosis. Less or no impact on income of retired participants. Spend of savings, selling of possessions, altering usual activities and enjoyment of life to cope with loss of income<br>c. Occurrence of negative emotions such as regret, disappointment, and self-reproach could lead to coexisting health problems and other difficulties. Family stress/strife, breakdown of relationships/families were other psychosocial facing patients and carers<br>d. Return to work prematurely due to financial commitments. Concerns about job loss, employability, and lack of promotion. |
| Callanan<br>2012 [28] | Benefits and allowances that families may be entitled to claim | Com<br>menta<br>ry |  | x | x |  |  |  | a. Increased financial burden due to loss of income and increased costs for special diet, new clothing, heating, travel, and car parking<br>b. Financial support from state welfare benefit system were needed the most by people with limited or no income |
| Moffatt<br>2012 [21] | Impact of welfare rights advice services on the quality of life and wellbeing of people with cancer | Mixed | P<br>+<br>C |  | x | x |  |  | b. Welfare benefits helped offset additional costs associated with cancer<br>c. Receiving welfare benefits reduced levels of stress and anxiety related to financial difficulties. |
| Rogers<br>2012 [22] | Financial burden of having head and neck cancer, and its relation with health-related quality of life (HRQoL) | Quan | P | x | x |  |  |  | a. 54% patients experienced at least one moderate or large financial burden. Greater financial difficulty due to loss of income. Younger people were more likely to experience financial difficulty<br>b. 39% patients applied for benefits. Of those applied, 71% had received it. Patients in working age and men were more likely to apply for benefits |
|  |  |  |  | a | b | c | d | e |  |
| Rogers<br>2012 [23] | Need for financial benefits, the advice patients were given about benefits and financial matters, and the financial burden of the disease | Quan | P | x | x |  |  | x | a. 57% reported that they were suffering financial hardship due to change in income<br>b. 63% claimed that they need benefits. Unemployed (91%), part-time employed (71%), and those whose work was affected by cancer (75%) were more likely to need benefits.<br>e. Decreased HRQoL (53%) as a result of the financial impact |
| Gardiner<br>2013 [32] | Financial costs and the financial impact of caring for family members receiving palliative/end-of-life care | Sys<br>rev |  | x | x |  |  |  | Included results from Hanratty et al (2007) |
| Macmillan<br>Cancer<br>Support<br>2013 [43] | Financial impact cancer is having on people across the UK | Mixed | P<br>+<br>S | x |  |  |  |  | a. 83% people are financially affected. The household was on average £570 a month worse off. Key factors that negatively influenced the severity of financial hardship were younger age (<60 years old), undergone chemotherapy and/or surgery, self-employed or part-time employed, and low income |
| McGarry<br>2013 [35] | Unmet supportive needs of people with breast cancer attending a London NHS Foundation Trust Hospital | Mixed | P | x | x | x |  |  | a. 17% of participants had concerns about finance (e.g., difficulties with rent and bills) due to inability to work or reducing of working hours<br>b. Support from the system was insufficient to meet patient's need and they had to depend on family for additional support<br>c. Financial concerns added to overall stress during treatment |
| Azzani<br>2015 [27] | Prevalence of perceived financial hardship and associated factors | Sys<br>rev |  | x | x |  |  |  | Included results from Rogers et al (2012) 'impact' |
| Moffatt<br>2015 [24] | Connections between cancer and employment; specifically, decisions, choice and constraints around returning to work or remaining outside the labour force | Qual | P | x | x | x |  |  | a. Affect household finances due to significant drop in income. Coping strategies are using savings, borrowing cash, cut on household expenditure, and selling property<br>b. Claiming welfare, even for a cancer-related illness, is stigmatising.<br>c. It was stressful due to concern over the impact of cancer on financial situation, future employment prospects, and families' life |
| Pelletier<br>2015 [36] | Family financial burden in childhood cancer | Sys<br>rev |  | x | x | x |  |  | Included results from Eiser et al (2006) |
|  |  |  |  | a | b | c | d | e |  |
| Young<br>Lives vs<br>Cancer<br>2016 [45] | Additional costs facing young cancer patients and their families; how a cancer diagnosis is disrupting the employment and income; emotional impact of the financial burden of cancer | Mixed | P<br>+<br>C | x | x | x |  |  | a. Parents spent £600 extra per month during active treatment of their children. For young patients, it was £360/month. Great financial pressure: 61% had built up debt and 17% borrowed over £5,000<br>b. Forms to apply for Disability Living Allowance and Personal Independence Payment was long and stressful to complete, and patients often required help to fill out (84%)<br>c. 76% of parents and 54% of young people reported additional stress and anxiety while managing their finances during treatment |
| Macmillan<br>Cancer<br>Support<br>2017 [42] | Financial impact of cancer |  | P<br>+<br>S | x |  | x |  | x | a. 39% of people with cancer have used savings, sold assets or borrowed to cover the costs or the loss of income caused by their diagnosis. 30% carers reported that their income or household finances were affected by caring<br>c. 53% reported feeling more anxious or stressed. 37% said it had made them feel more isolated or alone<br>e. Negatively affected quality of life (61%) |
| Watson<br>2019 [38] | Care experiences and supportive care needs | Quan | P | x |  |  |  |  | a. Negatively impacted on day-to-day financial situation (51%) |
| Flaum<br>2020 [31] | Financial toxicity (FT) and financial burden | Quan | P | x |  |  |  |  | a. Prevalence of FT among surveyed cancer patient was reported at 20% |
| Zhu<br>2020 [39] | Cancer survivors' experiences with financial toxicity | Sys<br>rev |  | x |  | x | x |  | Included results from Amir et al (2012) |
| Lu<br>2021 [25] | Association between levels of financial stress and cancer-related fatigue (CRF) | Quan | S | | | x | | | c. 11.% survivors reported both pre- and post-diagnosis financial stress (cumulative stress). Survivors with cumulative financial stress exposure were significantly more likely to have CRF (OR = 4.58, 95% CI 3.30-6.35, $p < 0.001$), compared with those without financial stress. |
| Fitch<br>2022 [30] | Cancer-related financial toxicity or burden | Sys<br>rev |  | x | x | x | x |  | Included results from Moffatt et al (2010), Moffatt et al (2012), and Amir et al (2012) |

*Study design – Mixed: Mixed methods, Qual: Qualitative research, Quan: Quantitative data, Rev: Narrative review, Sys rev: Systematic review* *Pop: Studies population – C: Carers/family members, P: Patients, S: Survivors*

#### Impact on financial well-being

24/29 studies reported that the patients, survivors, and/or carers faced severe financial problems following their cancer diagnosis [17, 19, 20, 22–24, 26–33, 35, 36, 38–45]. These problems manifested in varied forms such as being in debt [26, 29], difficulties paying rent/bills/mortgage [26, 35, 38], needing financial help [24, 33, 42], spending savings [24, 40, 42], selling possessions [24, 40, 42], altering usual activities and enjoyment of life to cope [24, 40]. The two main reasons leading to such situations were loss of income (e.g., due to needing to stop working or reduce working hours) and additional direct non-medical costs (e.g., special diet, heating, travel, and car parking).

Only 2/24 studies quantified the loss of income and extra expenditure. Studies from Macmillan reported that households, on average, were £570 worse off per month following a diagnosis of cancer [43]. Young Lives vs Cancer studies reported that parents spent £600 extra per month during active treatment of their children while young cancer patients spent £360 extra per month [45]. Likewise, few studies (5/24) reported the disparities among sociodemographic groups. These studies reported that financial impact was more severe for those of working age, especially self-employed or part-time employed [19, 22, 43]; lone parents [20]; and among those who belonged to minority ethnic groups [33].

#### Impact of benefit/welfare system

17/29 studies detailed the experiences that were reported by patients, survivors, and/or carers regarding the benefit/welfare system [17–19, 21–24, 26–28, 30, 32, 33, 35, 36, 41, 45]. Financial burden resulted in cancer patients applying for benefits such as attendance allowance, disability attendance allowance (DLA), and/or personal independence payment (PIP) [22, 33, 45] even though there was stigma associated with applying [18, 24]. Studies also reported patients’ dissatisfaction towards the benefit system - they complained that the application process was complicated and lengthy [26, 41, 45], the benefits that they received were inadequate [35], and there was an overall lack of information about benefit entitlements [18, 19]. All these issues added to the stress felt by patients (more details in next sub-section). 2/17 studies reported that patients who did not qualify for state benefits (their applications were denied) had to rely on grants from charitable organisations or support from family/friends [17, 22] though these two studies were published in 1982 and 2012.

#### Impact on mental health

16/29 studies reported how financial burden and struggles with obtaining benefits affected the mental health of patients, survivors, and/or carers [19–21, 24, 25, 29, 30, 35–37, 39–42, 44, 45]. The most common aspect reported was ‘worry about money’ which led to additional stress [19, 24, 35, 42, 45] and, as quoted, ‘if you’ve got money worries it brings you down a little bit further’ [19]. Lone parents were more likely to report money worries [20, 41]. Occurrence of negative emotions such as regret, disappointment, and self-reproach about their financial situation was viewed as leading to coexisting health problems and other difficulties [40]. Family stress/strife, breakdown of relationships/families were other significant psychosocial challenges facing patients and carers [40].

#### Impact on employment during and after treatment

5/29 studies reported the impact of financial burden on employment of patients, survivors, and/or carers during and after treatment [26, 30, 34, 39, 40]. All these studies reported that patients had to return to work prematurely due to financial pressure as a result of being off work.

#### Impact on health-related quality of life

Only 2/29 studies reported the impact of financial burden on the HRQoL of patients, survivors, and/or carers [23, 42]. Macmillan’s study reported that the HRQoL of 61% of patients was negatively affected though the validity of the method of measuring HRQoL was unclear [42]. Rogers et al (2012) reported that 53% of patients who suffered financially had decreased HRQoL as measured by the University of Washington Quality of Life questionnaire (UWQoL) [23].

## DISCUSSION

### Nature, extent and distribution of studies

No study in the UK has investigated FT as this term is commonly understood. Flaum et al (2020) was the only study with the explicit objective of investigating financial burden and FT. According to the authors, it was the first study of FT among UK cancer patients and it reported a prevalence of 20% of cancer patients who experienced FT [31]. However, the definition of FT in this study did not follow international practice. Financial burden was identified based on only one question (Q28) from the EORTC QLQ-C30 while FT was defined as greater financial burden at follow-up survey [31]. Objective financial burden and subjective financial distress were not clearly delineated in any study. As a result, we needed to adjust and broaden the inclusion criteria of the scoping review in order to include studies that reported any aspect of FT.

The significant increase in the number of publications on the subject in recent years reflects a growth of interest in the issue of FT within the field of cancer research. These publications came mostly from authors in academia though there were contributions from charity organisations and/or hospital Trusts. Indeed, charity organisations have published their own reports about the financial impact of cancer. These reports appeared to indicate a stronger presence of a wide variety of aspects related to financial impact than the peer-reviewed articles which tended to focus on only one aspect. While the voluntary sector has spearheaded research in this area, collaboration between the voluntary sector and academics may in the future help bring additional rigor to such studies and give a greater degree of credibility to their results.

Regarding geographical coverage, only one third of included studies provided UK-wide data. However, potential differences between England, Scotland, Wales, and Northern Ireland were not studied, despite the fact that these countries face distinct economic environments and organize health and social care differently [47]. This should be a consideration for further research.

### Methods of studies: Design, participants, exposure, and outcomes

Most common study designs were applied in included studies, including mixed-methods, a cross-sectional study, secondary data analysis, qualitative research, and a systematic review/review. However, a key limitation is that all quantitative studies used retrospective data. There has not been a study in the UK that has used prospective data to investigate the issue of FT among cancer patients. A prospective cohort study following patients from the point of diagnosis to finish initial treatment would provide invaluable insights to the causes and effects of FT on cancer patients. Such studies may be less likely to be subject to recall bias as well as providing the opportunity to study the relationship between FT and cancer as treatment progresses and/or economic conditions change.

The most studied population was cancer patients. Studies have paid some attention to survivors and carers/family members though they tended to be studied separately. Future research should assess the FT situation from the perspective of all key parties (i.e., patients, survivors, carers/family members) as well as the views of other stakeholders such as the community and voluntary sector. The involvement of one or more charity organisations in the recruitment process and the associated larger sample sizes in these studies points to the importance of adopting a collaborative approach in future research.

The validated questionnaire to investigate FT, COST, has not been used in any UK studies. Authors often used bespoke questionnaire or generic instrument which was not specialised for the issue of FT. It is recommended that future research should use COST to improve research rigour and facilitate the comparison of results with similar research around the world.

### Key findings of studies - dominant areas of research amongst studies

The majority of studies focused on describing objective financial burden and its material impact on the financial well-being of cancer patients, survivors, and/or carers/family members. Subjective financial distress, especially its psychosocial effects, is under-researched. There is a need too to give research attention to investigating disparities between different sociodemographic groups. The review found that studies are sparse regarding the causes of financial stresses and strain. Whilst some FT-related outcomes were investigated (e.g., impact on financial well-being, mental health, employment, HRQoL), there is a need to assess FT using psychometrically validated instruments. These critical gaps for future research need to be addressed in order to plan person-centred service responses for patients who encounter FT.

### Strengths and limitations

This scoping review provides a comprehensive and thorough review of available literature about financial toxicity among cancer patients, survivors, and their families in the UK. As a first step, the scoping review identified key research gaps and suggested priorities for future research. Our comprehensive and systematic approach to identification, selection, data charting and analysis followed the rigorous methodological framework set out by Arksey & O’Malley (2007) and Levac et al (2010) [14, 15] and PRISMA guidance for the conduct and reporting of scoping reviews [16]. We plan to carry out (optional) stage 6 of the methodological framework (i.e., consultation exercise) in the months ahead.

## CONCLUSIONS

There exists a paucity of research on financial toxicity among cancer patients, survivors and their families in the UK. Current evidence is ad hoc and at times anecdotal with studies using different definitions, methods and studying often only small parts of the overall issues. Nevertheless, that FT exists in the UK is evident. A comprehensive study designed to provide a better understanding about the nature and extent of the problem, disparities in experience (among different sociodemographic groups and types of cancer), the impacts of FT on outcomes, and potential solutions to alleviate FT and related problems is urgently needed.

## Data Availability

All data produced in the present work are contained in the manuscript

## Additional information

## Acknowledgements

The authors would like to thank Mr Richard Fallis who is the subject librarian of Medicine, Dentistry & Biomedical Sciences at Queen’s University Belfast for his advice on the development of the search strategy.

## Authors’ contributions

TTN, CON, and MD conceived and designed the scoping review. TTN and THT participated in the study selection and data extraction. All authors contributed to the interpretation of the findings. TTN wrote the first draft and prepared the manuscript. MD and CON provided supervisory support and reviewed this paper. All authors contributed to the revision of the manuscript and approved the final version of the review.

## Ethics approval and consent to participate

Not applicable

## Consent for publication

Not applicable

## Data availability

Search strategies needed to replicate the study are included in Table S1, Supplementary information.

## Supplementary information

Table S1 Detailed search strategies for all databases/search engine

Table S2 Detailed inclusion and exclusion criteria

Appendix 1 PRISMA extension for scoping reviews (PRISMA-ScR) checklist

Appendix 2 List of excluded reviews and justification for the exclusions

