## Supplementary information for "Financial toxicity among cancer patients, survivors, and their families in the United Kingdom: a scoping review"

#### Table S1: Detailed search strategies for all databases/search engine

##### a) EMBASE (via Ovid)

| |
|---|
| 1. malignant neoplasms/ or carcinoma.mp. [mp=title, abstract, heading word, drug trade name, original title, device manufacturer, drug manufacturer, device trade name, keyword heading word, floating subheading word, candidate term word]<br>2. (cancer* or carcinoma* or malignan* or neoplasm* or tumo?r*).ti,ab.<br>3. (financial* adj9 (hardship* or stress* or burden* or distress* or strain* or toxicity* or catastrophic* or consequence* or impact*)).ti,ab. 4. physical examination/ or palpation/<br>4. (economic* adj5 (hardship* or burden* or strain* or consequence* or impact*)).ti,ab. 6. "clinical breast examination*".ti,ab.<br>5. 1 or 2<br>6. 3 or 4<br>7. 5 and 6<br>8. exp United Kingdom/<br>9. 7 and 8<br>10. limit 9 to english language |

##### b) Google Scholar

| |
|---|
| "financial" AND "cancer" AND ("united kingdom" OR "england" OR "scotland" OR "wales" OR "northern ireland") |

##### c) MEDLINE (via Ovid)

| |
|---|
| 1. neoplasms/ or carcinoma.mp. [mp=title, book title, abstract, original title, name of substance word, subject heading word, floating sub-heading word, keyword heading word, organism supplementary concept word, protocol supplementary concept word, rare disease supplementary concept word, unique identifier, synonyms]<br>2. (cancer* or carcinoma* or malignan* or neoplasm* or tumo?r*).ti,ab.<br>3. (financial* adj9 (hardship* or stress* or burden* or distress* or strain* or toxicity* or catastrophic* or consequence* or impact*)).ti,ab.<br>4. (economic* adj5 (hardship* or burden* or strain* or consequence* or impact*)).ti,ab.<br>5. 1 or 2<br>6. 3 or 4<br>7. 5 and 6<br>8. exp United Kingdom/<br>9. 7 and 8<br>10. limit 9 to english language |

**d) Scopus**

| |
|---|
| ( ( TITLE-ABS-KEY ( financial* W/9 ( hardship* OR stress* OR burden* OR distress* OR strain* OR toxicity* OR catastrophic* OR consequence* OR impact* ) ) OR TITLE-ABS-KEY ( economic* W/5 ( hardship* OR burden* OR strain* OR consequence* OR impact* ) ) ) AND TITLE-ABS-KEY ( cancer* OR carcinoma* OR malignan* OR neoplasm* OR tumo*r* ) AND TITLE-ABS-KEY ( "united kingdom" ) AND LANGUAGE ( english ) ) |

**e) Web of Science**

| | |
|---|---|
| #1 | cancer* or carcinoma* or malignan* or neoplasm* or tumo$r* (Topic) |
| #2 | TS=((financial* near/9 (hardship* or stress* or burden* or distress* or strain* or toxicity* or catastrophic* or consequence* or impact*))) |
| #3 | TS=((economic* near/5 (hardship* or burden* or strain* or consequence* or impact*))) |
| #4 | #2 OR #3 |
| #5 | #4 AND #1 |
| #6 | TS=(United Kingdom) |
| #7 | #5 AND #6 |
| #8 | (#5 AND #6) AND LA=(English) |

**Table S2: Detailed inclusion and exclusion criteria**

| Study | Inclusion criteria | Exclusion criteria |
|---|---|---|
| **Population** | ONLY those living in the United Kingdom (including England, Scotland, Wales, and Northern Ireland)<br>• Cancer patients (those who are under treatment, including patients with initial treatment after diagnosis and people with on-going treatment* for advanced cancer)<br>• Cancer survivors (those who finished initial**/hospital treatment). Other terms: cancer-free survivors, disease-free survivors<br>• Family members of cancer patients/survivors (whether giving care to patients/survivors or not). Other terms: carer, caregiver<br><br>**NOTE**<br>** On-going treatment is different from follow-up treatment. The latter is for cancer-free survivors and normally includes hormone therapy and/or periodic check-up (last 5-10 years)*<br>*** Initial/hospital treatment: Treatment provided after diagnosis and before discharge (cancer-free). It often lasts up to 9-12 months, depends on type of cancer and stage of cancer at diagnosis* | • Non-human subjects<br>• Cancer patients/survivors or carers reside in any countries rather than the UK |
| **Exposure (the issue)** | • Financial toxicity (refers to both the **objective financial burden** and **subjective financial distress**) experienced by the population of interest***<br>• Other terms: financial hardship/stress/burden/distress/strain/catastrophic, economic burden/consequence/impact/strain<br><br>**NOTE**<br>*** Patient perspective: the review only focuses on the issue (financial toxicity) from the population of interest's perspective. Studies report the patient-level data of the issue will be included | • Studies report the economic burden of the disease, financial impact of the treatment, drug etc. from the society or payer perspective<br>• Studies report the cost of treatment, drug without discuss the experience of patients/survivors bearing that cost<br>• Cost-effectiveness analysis of treatment/drug |

| Study | Inclusion criteria | Exclusion criteria |
|---|---|---|
| **Outcomes** | **Primary main outcome**<br>• Financial well-being<br>• Health-related quality of life<br>• Additional mental health conditions such as depression and anxiety<br>**Secondary main outcomes**<br>• Benefit/welfare benefit<br>• Counselling service<br>• Any other support with purpose to ease FT | • Outcomes not listed |
| **Study context** | • United Kingdom (including England, Scotland, Wales, and Northern Ireland) | • Any other countries |
| **Publication type** | • Published at anytime<br>• English language only<br>• Any types, including grey literature | • None (no time restriction)<br>• Non-English articles<br>• Publications that do not have an available full-text version or are duplications of other publications |

**Appendix 1: Preferred Reporting Items for Systematic reviews and Meta-Analyses extension for Scoping Reviews (PRISMA-ScR) Checklist**

| SECTION | ITEM | PRISMA-ScR CHECKLIST ITEM | REPORTED ON PAGE # |
|---|---|---|---|
| **TITLE** | | | |
| Title | 1 | Identify the report as a scoping review. | Page 1, line 1,2 |
| **ABSTRACT** | | | |
| Structured summary | 2 | Provide a structured summary that includes (as applicable): background, objectives, eligibility criteria, sources of evidence, charting methods, results, and conclusions that relate to the review questions and objectives. | Page 2, line 20-34 |
| **INTRODUCTION** | | | |
| Rationale | 3 | Describe the rationale for the review in the context of what is already known. Explain why the review questions/objectives lend themselves to a scoping review approach. | Page 3, 4. Line 39-81 |
| Objectives | 4 | Provide an explicit statement of the questions and objectives being addressed with reference to their key elements (e.g., population or participants, concepts, and context) or other relevant key elements used to conceptualize the review questions and/or objectives. | Page 4, line 83-86 |
| **METHODS** | | | |
| Protocol and registration | 5 | Indicate whether a review protocol exists; state if and where it can be accessed (e.g., a Web address); and if available, provide registration information, including the registration number. | Review protocol exists but was not registered |
| Eligibility criteria | 6 | Specify characteristics of the sources of evidence used as eligibility criteria (e.g., years considered, language, and publication status), and provide a rationale. | Page 5, line 106-107. Page 6, line 121-130 |
| Information sources* | 7 | Describe all information sources in the search (e.g., databases with dates of coverage and contact with authors to identify additional sources), as well as the date the most recent search was executed. | Page 5, 6. Line 103-118 |
| Search | 8 | Present the full electronic search strategy for at least 1 database, including any limits used, such that it could be repeated. | Table S1, supplementary information |
| Selection of sources of evidence† | 9 | State the process for selecting sources of evidence (i.e., screening and eligibility) included in the scoping review. | Page 6, line 112-118 |
| Data charting process‡ | 10 | Describe the methods of charting data from the included sources of evidence (e.g., calibrated forms or forms that have been tested by the team before their use, and whether data charting was done independently or in duplicate) and any processes for obtaining and confirming data from investigators. | Page 7, line 138-146 |
| Data items | 11 | List and define all variables for which data were sought and any assumptions and simplifications made | Page 7, line 142-146 |

| SECTION | ITEM | PRISMA-ScR CHECKLIST ITEM | REPORTED ON PAGE # |
|---|---|---|---|
| Critical appraisal of individual sources of evidence§ | 12 | If done, provide a rationale for conducting a critical appraisal of included sources of evidence; describe the methods used and how this information was used in any data synthesis (if appropriate). | Not applicable |
| Synthesis of results | 13 | Describe the methods of handling and summarizing the data that were charted. | Page 7, line 148-153 |
| **RESULTS** | | | |
| Selection of sources of evidence | 14 | Give numbers of sources of evidence screened, assessed for eligibility, and included in the review, with reasons for exclusions at each stage, ideally using a flow diagram. | Page 7, line 156-163<br>Figure 1 |
| Characteristics of sources of evidence | 15 | For each source of evidence, present characteristics for which data were charted and provide the citations. | Page 8, 9. Line 164-214<br>Table 1 |
| Critical appraisal within sources of evidence | 16 | If done, present data on critical appraisal of included sources of evidence (see item 12). | Not applicable |
| Results of individual sources of evidence | 17 | For each included source of evidence, present the relevant data that were charted that relate to the review questions and objectives. | Table 2, 3 |
| Synthesis of results | 18 | Summarize and/or present the charting results as they relate to the review questions and objectives. | Page 9-12. Line 216-274 |
| **DISCUSSION** | | | |
| Summary of evidence | 19 | Summarize the main results (including an overview of concepts, themes, and types of evidence available), link to the review questions and objectives, and consider the relevance to key groups. | Page 12-14. Line 275-335 |
| Limitations | 20 | Discuss the limitations of the scoping review process. | Page 15, line 338-345 |
| Conclusions | 21 | Provide a general interpretation of the results with respect to the review questions and objectives, as well as potential implications and/or next steps. | Page 15, line 348-354 |
| **FUNDING** | | | |
| Funding | 22 | Describe sources of funding for the included sources of evidence, as well as sources of funding for the scoping review. Describe the role of the funders of the scoping review. | Page 316, line 371-373 |

JBI = Joanna Briggs Institute; PRISMA-ScR = Preferred Reporting Items for Systematic reviews and Meta-Analyses extension for Scoping Reviews.
* Where *sources of evidence* (see second footnote) are compiled from, such as bibliographic databases, social media platforms, and Web sites.
† A more inclusive/heterogeneous term used to account for the different types of evidence or data sources (e.g., quantitative and/or qualitative research, expert opinion, and policy documents) that may be eligible in a scoping review as opposed to only studies. This is not to be confused with *information sources* (see first footnote).
‡ The frameworks by Arksey and O'Malley (6) and Levac and colleagues (7) and the JBI guidance (4, 5) refer to the process of data extraction in a scoping review as data charting.
§ The process of systematically examining research evidence to assess its validity, results, and relevance before using it to inform a decision. This term is used for items 12 and 19 instead of "risk of bias" (which is more applicable to systematic reviews of interventions) to include and acknowledge the various sources of evidence that may be used in a scoping review (e.g., quantitative and/or qualitative research, expert opinion, and policy document).

#### Appendix 2: List of excluded reviews and justification for the exclusions

This list includes 30 articles that were excluded after full-text review with justification for the exclusion of each. The list does not contain articles excluded due to duplicate or during the title and abstract screening.

**Ineligible population:** One review was excluded

1. Cook NS, Landskroner K, Shah B, Walda S, Weiss O, Pallapotu V. Identification of Patient Needs and Preferences in Pigmented Villonodular Synovitis (PVNS) Using a Qualitative Online Bulletin Board Study. Advances in therapy. 2020;37(6):2813-28.

**Ineligible exposure (no data on FT):** Ten reviews were excluded

**Ineligible outcomes:** No review was excluded

**Wrong setting:** Five reviews were excluded

**No full-text available:** Fourteen reviews were excluded

17. Brearley SG, Craven O, Wilson B, Brunton L, Molassiotis A. Gastro-intestinal cancer patients: How they perceive and cope with disease and treatment-related symptoms over a 12-month period. European Journal of Cancer, Supplement. 2009;7(2-3):234.
18. Choon-Quinones M, Zelei T, Barnett M, Keown P, Durie B, Kalo Z, et al. Exploring the true cost of multiple myeloma. American Journal of Hematology. 2020;95(SUPPL 1):S8-S9.
19. Corney R, Swinglehurst J, Brett-Smith D. The impact of breast cancer on couple and family relationships of young women. Psycho-Oncology. 2011;20(SUPPL. 2):93-4.
20. Cox T, MacLennan S, Scott S. Cancer survivorship and working life. Psycho-Oncology. 2014;23(SUPPL. 3):355-6.
21. Higginson IJ, Gomes B, Calanzani N, Gao W, Bausewein C, Daveson BA, et al. Factors associated with the priorities for treatment and care if faced with advanced cancer across seven european countries. Palliative Medicine. 2012;26(4):410.
22. Holmes L, Addington-Hall J, Grande G, Payne S, Seymour J, Hanratty B. Transitions between care settings in the last year of life for people living with heart failure, stroke and lung cancer: A qualitative study. Palliative Medicine. 2010;24(4 SUPPL. 1):S190.
23. Hyman J, Lucas E. Community cancer programs network-bringing cancer care closer to home. Asia-Pacific Journal of Clinical Oncology. 2014;10(SUPPL. 9):27.
24. Mowbray M, Fraser S, Hancock E, Scorgie C. Melanoma patient support-what does your patient want? Melanoma Research. 2016;26(Supplement 1):e59.

25. Nelson D, Pascal J, Kane R, McGonagle I, Kenny A, Dickson-Swift V, et al. The psychosocial support needs of people affected by cancer: A comparative study of patient and carer experience in a rural setting. Psycho-Oncology. 2017;26(Supplement 3):80.
26. Palmer SK, Collie J. Transplant experience for post-transplant patients who travel for follow-up care. Bone Marrow Transplantation. 2011;46(SUPPL. 1):S448.
27. Reaney M, Eek D, Ascoytia C, Scrabis L, Halling K, Black P, et al. Similarities and differences between symptoms and impacts of ovarian cancer as reported by the patients and their caregivers. European Journal of Cancer. 2015;51(SUPPL. 3):S248.
28. Scanlon K, Tompkins C, Ream E, Armes J, Harding S. Challenging the concept of self management: Ethnic minority women's experiences of early breast cancer survivorship. Psycho-Oncology. 2013;22(SUPPL. 3):41.
29. Tanguay JS, Long J, Knoyle DM, Gibson A. Maximising financial aid for patients in a thoracic oncology clinic in merthyr tydfil. The effect of a clinic-based Welfare Rights Officer. Lung Cancer. 2013;79(SUPPL. 1):S42.
30. Younger E, Husson O, Desar IME, Young R, Leahy MG, Oosten A, et al. Health-related quality of life in patients with advanced soft tissue sarcomas treated with chemotherapy: The HOLISTIC study. Annals of Oncology. 2019;30(Supplement 5):v709.

**Appendix 3: Data charting template (used in Covidence)**

#### 1. GENERAL INFORMATION

**Study ID**

**Lead author -** *Surname only (e.g., Amir)*

**Year of publication**

**Title**

*Title of paper / abstract / report that data are extracted from*

**Type of publication**

- (•) Peer-reviewed article
- ( ) Grey literature (e.g., report)

**Geographic coverage**

*If UK-wide, tick all options. If study is a review/sys review that have more locations, note in the 'other' field*

- ☐ England
- ☐ Scotland
- ☐ Wales
- ☐ Northern Ireland
- ☐ Other

**Authors' affiliation**

*Note where the authors work (and their position if available). e.g., university, hospital*

#### 2. CHARACTERISTICS OF INCLUDED STUDIES

##### a) Methods

**Objectives/research questions**

**Study design**

- ( ) Randomised controlled trial
- ( ) Cohort study
- ( ) Cross sectional study
- ( ) Systematic review
- ( ) Qualitative research
- ( ) Narrative review
- ( ) Scoping review
- ( ) Mixed methods
- ( ) Others

#### b) Participants (skip in case of rev/sys rev)

**Studied population**

- ☐ Patients
- ☐ Survivors
- ☐ Carers/family members
- ☐ Other

**Type of cancer (that patients/survivors have/had)**

**Total number of participants**

**Participants' characteristics**

*(e.g., age, socio-economic groups) This is not the data from table 1 but the inclusion criteria for participants*

**Method of recruitment of participants**

- ☐ Phone (random dial)
- ☐ Mail (postal study)
- ☐ Online
- ☐ In-person recruitment (at hospital, support centre, etc.)
- ☐ Other

**Setting of the recruitment (Specify)**

*E.g., hospital, cancer registry, cancer forum, support centre etc.*

#### c) Included studies (specifically for rev/sys rev)

**How many studies from UK were included in the rev/sys rev?**

*Number only*

**Which studies were included?**

*Author, year, title. Note is the original study was in this scoping review or not (e.g., Amir 2012 xyz, IN/NOT IN).*

#### d) Exposure (Financial Toxicity)

**Used terms**

*The terms were used to describe the problem (e.g., financial hardship, stress)*

**Exposure description**

*Give a detailed description of the studied problems here*

**Tools used to measure FT**
*If validated tool, note the name (e.g., COST, EUQOL). Otherwise, record the questions & answer options*

**Subjective financial distress sub-category**
*What aspect(s) did the included study look at?*

- ☐ Material
- ☐ Psychosocial
- ☐ Behavioral

#### e) Outcomes

**What are the outcomes?**

- ☐ Financial well-being/situation
- ☐ Health-related quality of life
- ☐ Additional mental health conditions such as depression and anxiety
- ☐ Employment (work/retirement)
- ☐ Benefit/welfare
- ☐ Any other support with purpose to ease FT
- ☐ Others

### 3. RESULTS

**Key findings**
Summary the key findings of the study

**Future research/recommendations**
*This is normally mentioned in the limitation, end of discusison section*

**Conclusions of the research**

---

**Final thought**
*Anything you think that is important but was not captured by this form*
